# Dynamic coupling between the COVID epidemic timeline and the behavioral response to PAUSE in New York State counties

**DOI:** 10.1101/2020.07.08.20148981

**Authors:** Anca Rǎdulescu, Shelah Ballard, Johnathan Linton, Kaitlyn Gonzalez

## Abstract

By many expert opinions, the COVID 19 epidemic is still much in its developing stages. While awaiting for effective clinical answers for the outbreak, the best approach remains that of social distancing. This raises a few crucial questions related to the extent to which social distancing measures were (1) well timed, (2) necessary and (3) efficient. By investigating correlations between epidemic measures such as daily infection rate, and social mobility measures (as reported by Apple and Google), our study aims to establish whether data identifies social distancing measures as a primary player in controlling the outbreak in New York State.

## 1 Introduction

Early epidemic development is typically characterized by an initial period with high growth rate, after which the growth rate subsides to a slower curve, reaches a peak, and eventually transitions to a decreasing trend (as the epidemic is dying out). It is common that this first “wave” of the outbreak is followed by others, the timing, size and dynamics of which depend on a variety of epidemic factors (whether the virus confers immunity or not, whether treatment or prevention plans are available), demographic factors (population density and birth rate) and mitigation factors (presence and efficiency of social distancing measures).

The first confirmed infections with COVID-19 were identified in the US at the end of January 2020. Since then, the outbreak has spread dramatically to all US states and territories. The first response to the overwhelming pandemic seemingly attempted to compensate for the clinical unpreparedness (absence of a treatment or prevention plan, notorious limitations in testing, capacity of the health care resources). Since efficient containment failed, the response consisted of a system of mitigation measures based around social isolation (travel bans, shut down of non-essential businesses, prohibitions of large gatherings). The general population was asked to either quarantine or observe social distancing, depending upon suspicion of active symptoms, and upon the gravity of the local situation.

There have been, however, dramatic differences in the timeline and magnitude of the epidemic across different states. It is likely that these differences are based on a combination of inherent factors, from timing of the first infection (earlier states were caught unprepared), to population density and intrinsic social dynamics, but also on state-centralized factors such as timing and efficiency of state-wide mandated directives (travel bans, social distancing, timing and availability of testing). While some states have transcended the first epidemic peak and are currently experiencing a respite in daily infection rates, other states are experiencing dramatic surges in numbers. It is not clear whether this is a refueling of the first infection wave, or the start of a second wave altogether; either way, these latter states are facing the possibility of instating new lockdown measures (or strengthening existing ones), in am attempt to avoid exceeding medical resources and their health care capacities (which was the case in New York State in March and April 2020).

New York State had undoubtedly the most dramatic early surge (peaking at over 11K infections per day in mid April), but has since been seemingly able to control the outbreak (with consistently under 1.5K infections per day since June 5th, and generally still on a decreasing path). It has been hypothesized that this is the effect of a strict and efficient system of government mandated lockdown and social distancing measures (known as PAUSE), in combination with a carefully staggered reopening schedule. In our study we aim to test this hypothesis across New York counties, based a correlation analysis between the epidemic data on one hand (assessing the outbreak size and dynamics) and the social mobility data on the other hand (assessing the behavioral efficiency of the state mandated measures).

In terms of policy, New York State’s response to the pandemic came in a few stages, both in terms of testing and of mandated social measures. In terms of social dynamics restrictions, among the first to close in the New York State were university campuses and schools [1, 2], followed by restaurants, gyms and other entertainment venues [3, 4]. Along the month of March, gathering of gradually smaller sizes were banned, all eventually leading to the PAUSE directive that took effects on March 22nd and shut down state-wide all non-essential activity [5, 6]. PAUSE was fully in effect until the start of June, when the process of staggered reopening started, with stages tailored specifically to the situation in each county or region.

By many expert opinions, the COVID 19 epidemic is still much in its developing stages. While awaiting for effective clinical answers for the outbreak, the best approach remains that of social distancing. This raises a few crucial questions related to the extent to which social distancing measures were (1) well timed, (2) necessary and (3) efficient. Our study aims to establish whether data identifies social distancing measures as a primary player in controlling the outbreak in New York State.

We will investigate how quickly and to what extent people had responded to PAUSE in different counties, and how strictly they have been observing social distancing measures since then. We will allow data to inform which social activities had the highest correlations with the surge in infections, but also with the curbing in infection rates. Our final goal is to understand (1) to which extent the magnitude of the epidemic in each county modulated the efficiency of the social dynamics response to PAUSE measures; (2) to what extent the tightening and subsequent relaxation in social mobility successfully mitigated the epidemic effects. What makes the assessment of PAUSE efficiency particularly difficult is the absence of control data (we only have real infection data under PAUSE measures, but no estimates of infection counts in absence of social distancing measures). A notorious discussion point lies around the popular misconception that measures may have been exaggerated (put in perspective of the outbreak currently decreasing in New York State), when in fact it is the presence of measures that helped control the outbreak.

Establishing with high confidence whether and which social distancing measures are efficient is crucial in the context of forging optimal reopening strategies, planning a safe response to potential future waves. A lot of effort is currently being directed towards predictive epidemic models of COVID based on the existing data at hand. The answers that our study provides can inform such models, and generate new testable predictions.

The rest of the paper is organized as follows: in Section 2, we present our data-sets, the characteristics of our time series, and the measures we extracted from this data to assess the size of the epidemic (on one hand), and the efficiency of the state mandated lock down measures (on the other hand). We also describe the mathematical methods used for data processing and analysis, including the troubleshooting and alternative approaches that we had to design in order to best extract robust patterns from incomplete or imperfect data. In Section 3.1, we investigate separately the evolution of the outbreak across counties, since its first New York confirmed infection (March 2nd), until June 21st. In Section 3.2 we analyze the traffic patterns from January 13 until June 14th (as provided by Apple); in particular, we identify commonalities and differences between counties. In Section 3.3, we analyze mobility patterns tracking the number of people headed to different targets, as well as their residential time; we aim to understand the psycho-social drives underlying county specific, geography-specific, and week day specific trends. In Sections 3.4, 3.5 and 3.6, we calculate correlations between these data, which help us understand the subtle coupling between the epidemic dynamics on one hand, and social dynamics under lock down measures, on the other. In Section 4, we interpret our results, and infer potential cause-effect relationships between the epidemic timeline, and the efficiency of lock down.

## 2 Modeling methods

### 2.1 Data sources and processing

#### Epidemic data

Time series for total and daily numbers of confirmed infections, as well as for total and daily numbers of COVID 19 tests performed – broken down by New York State county – were accessed from March 2 (when the infection was first detected in New York) until June 21 (current date) from the New York Health Department data repository [8]. Population count and density measures for each county were found in the 2010 New York State census [9]. To track the evolving size of the outbreak, we used, for each county: (1) the number of confirmed cases; (2) the normalized number of cases (reported per 1000 individuals in each county); (3) incidence of positive daily tests versus total number of daily tests; (4) daily infection rates. Since the daily rates showed pronounced weekly oscillations (largely due to corresponding oscillations in reporting), we worked with smoothed time series obtained by using a seven day moving window average, without losing any of the long-term intrinsic epidemic trends (as shown in Figure 1).

**Figure 1:**
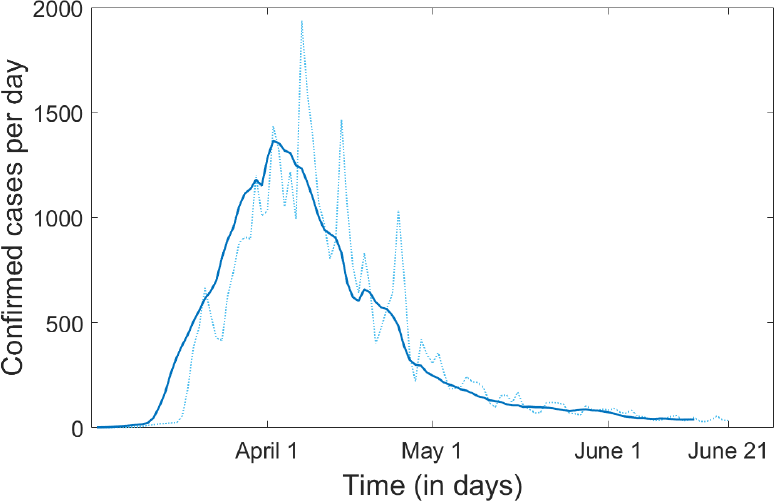
Example of time series showing the daily rate of confirmed infections, and its windowed smoothing for New York County. The raw data is represented as a dotted curve; the processed time series obtained by using a seven day average moving window is shown as a solid curve.

#### Traffic data

Time series describing traffic activity between January 13 and June 19 were made available in the public domain by Apple [10], based upon their daily numbers of hits for driving / walking / public transit directions requests. The accessible data reports the number of hits originating in each individual county as a percent of the baseline for that county, defined by the county-wide number of hits on January 13 (but not explicitly provided). While the county-specific baseline makes magnitude comparisons meaningless between different counties, one can still compare the trends in the time series (e.g., one county had a steeper drop than another).

Since the daily time series showed pronounced weekly oscillations, we obtained (as in the case of infection rates) smoothed time series, by using a seven day moving window average, as shown in Figure 2. One additional problem, which we will discuss later, is the absence of control time series. Since Apple did not make available any data from previous years, it is impossible to interpret the effect of seasonality of of annual events (such as increasing trends in traffic with the spring months, or specifically on Memorial weekend).

**Figure 2:**
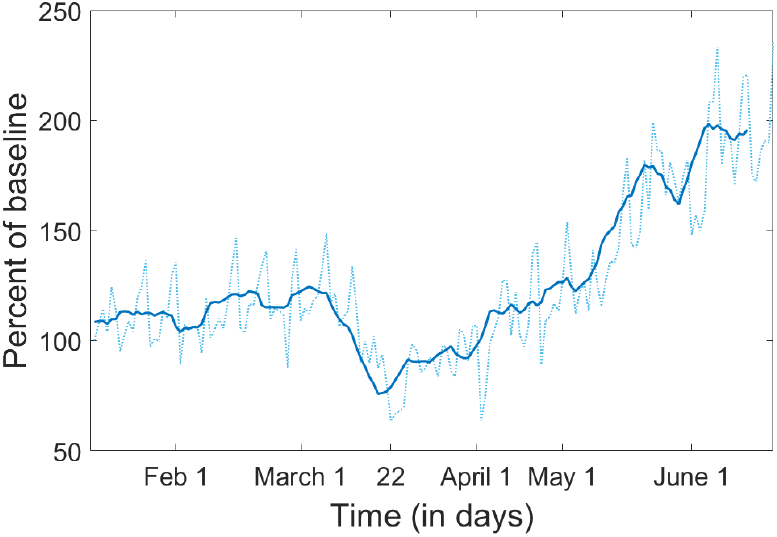
Example of driving time series and window smoothing for Orange county. The raw data is represented as a dotted curve; the processed time series obtained by using a seven day average moving window is shown as a solid curve.

Records were completely missing for two days (May 11 and 12) in the Apple traffic reports for all counties. For visual purposes only (when representing the time series), we filled these two values by taking the average of the previous and following week readings, before applying the moving window smoothing.

#### Mobility data

Google reports are based in each county on the number of daily searches for directions to five types of destinations (Retail/Recreation, Groceries/Pharmacy, Parks, Transit and Workspace), as well as on the daily amount of time spent at the residence. These are reported for each county with respect to a set of seven different (week day specific) baselines (not provided on the public domain), computed as the average of the activity on the specific week day for the five week period between January 3 and February 6. As for the traffic data, the county-specific baselines make magnitude comparisons meaningless, but this still allows the possibility to compare trends between counties. Unfortunately, reports are omitted for a days in which the county wide numbers did not meet the privacy threshhold. This effect was different in different counties (more pronounced in those with small population size) and for different targets (Parks were most affected, while Retail, Groceries and Workspace were least affected), and its prevalence changed along the timeline. This limited our analysis to the modalities for which there was sufficient data for an assessment.

An additional difficulty with this data set is the changing baseline, which compromises the longitudinal consistency of the time series. In order to address this problem, we processed the raw time series in two different ways. One was to consider, like in the case of infection rate and traffic time series, a seven window moving average (Figure 3). However, while this smoothes out the weekly oscillations in the time course and balances some of the baseline discrepancies, it is not a rigorous way to eliminate the dependency on the seven different baselines. Therefore, we also adopted in parallel a different approach, and calculated week day specific time series (e.g., “Sunday weekly series,” or “Monday weekly series”) in which all values are reported with respect to the common baseline for the specific day (Figure 4). Notice that, for Workspace mobility, the weekend time series appear a lot higher than the Monday-Friday ones. This does not mean that mobility to Work was higher during the weekend, but rather that weekend mobility shows a more shallow drop (since the baseline was already low). Similarly, the weekend Residential time series appear lower than the Monday-Friday ones, reflecting the higher weekend baseline (the typically higher number of residential time during weekends).

**Figure 3:**
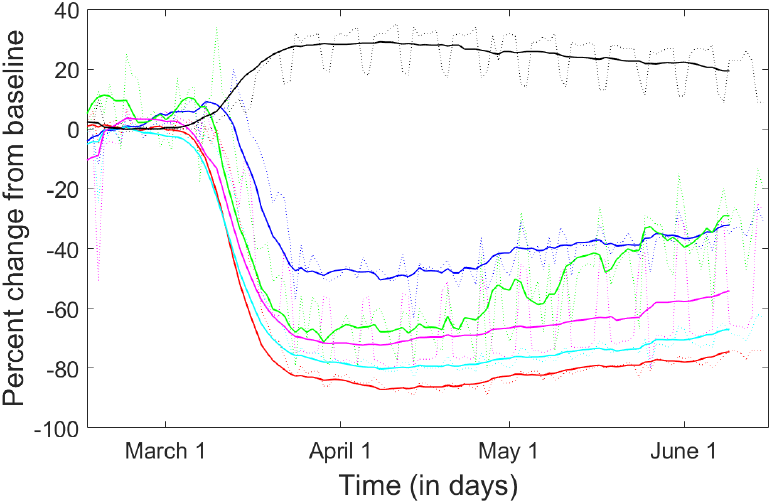
Example of mobility time series and window smoothing for New York County (i.e., Manhattan). The raw data is represented as a dotted curve: in red for Retail; in blue for Groceries; in green for Parks; in cyan for Transit; in pink for Workspace; in black for Residential. The processed time series obtained by using a seven day average moving window are shown as solid curves, with the same color coding.

**Figure 4:**
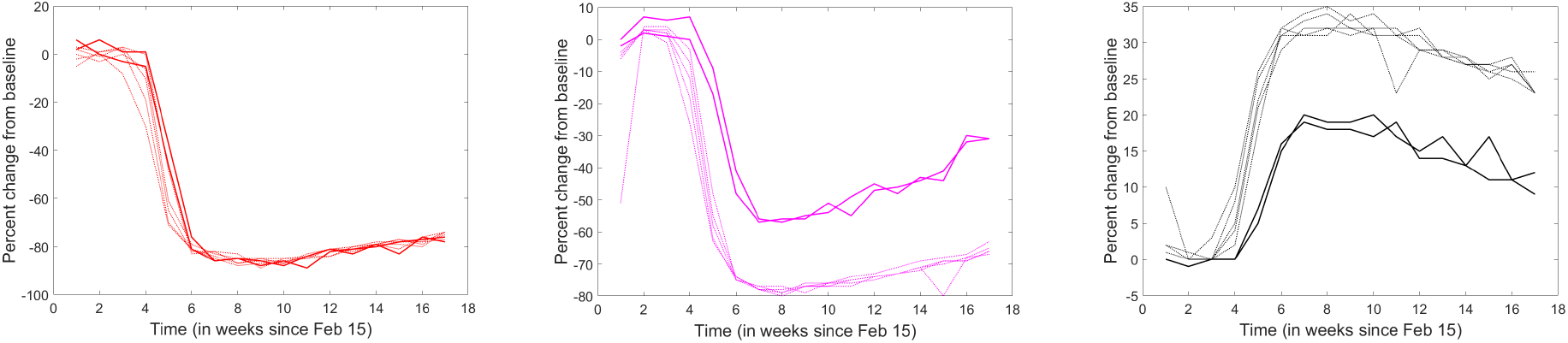
Example of weekly time series for **A**. Retail, **B**. Workspace and **C**. Residential mobility in New York County. The Monday-Friday time series are shown as dotted lines, and the weekend time series are shown as solid lines.

### 2.2 Correlation analysis

In order to measure the strength of the linear association between epidemic and traffic/mobility variables (say X and Y), we used a nonparametric measure, the Spearman rank correlation (the Pearson correlation coefficient between rank variables):

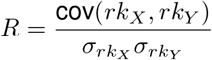

where *rk_X_* and *rk_Y_* are the rank variables corresponding to *X* and *Y*.

Spearman’s correlation assesses monotonic relationships (whether linear or not), with a positive value 0 *< R* ≤ 1 when X and Y tend to simultaneously increase, and a negative value − 1 ≤ *R <* 0 if Y tends to increase when X decreases. The Spearman correlation increases in magnitude as X and Y become closer to being perfectly monotone functions of each other. We used the Spearman correlation, because it is less sensitive than the Pearson correlation to strong outliers in the tails of both samples (since the outlier is limited to the value of its rank).

## 3 Results

### 3.1 Infection trends

Since the start of June, as the New York State has started its staged reopening, the state-wide number of confirmed COVID cases shows an almost asymptotic trend, with steadily decreasing daily rates (which was in fact one of the reopening benchmarks). While all counties with significant infection show the same slowing down growth to “capacity” in total infection counts, Figure 5a illustrates the wide variability in the sheer size among counties. New York City counties lead the first tier, followed within the same order of magnitude by suburban counties which were also among the early infection wave (Nassau, Suffolk, Westchester, Rockland, Orange). When normalizing the confirmed case count by the population size in each county, as reported on the 2010 census (Figure 5b), the order changes slightly (with Rockland and Westchester on top, and New York County less significantly affected, when accounting for its huge population size), but the primary infection hubs identified by this measure are the same as for the raw case counts.

**Figure 5:**
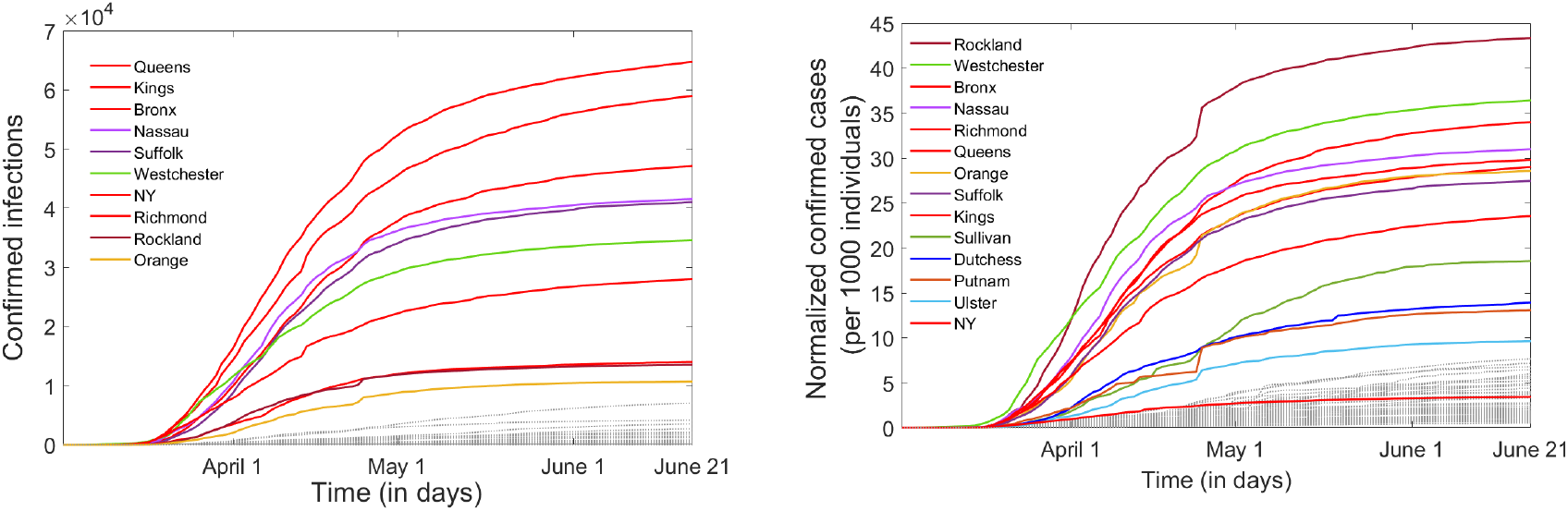
Infection time course in New York counties from May 2 until June 21. **A**. Total number of confirmed infections; **B**. Number of confirmed infections normalized by the county population size (reported per 1,000 individuals). The counties with the highest number of confirmed infections by June 21 are emphasized as solid curves, and labeled top to bottom in the legend. The rest of the counties, with lower infection counts, are shown as faded dotted lines, for comparison.

A more helpful representation of the epidemic dynamics is provided by the time series of the daily infection rates (i.e., the the daily rate of change in confirmed infections) shown in Figure 6. The rate-based representation better illustrates the rise and fall of the first epidemic wave in New York, and strongly supports the idea that the outbreak has been at least temporarily controlled. The object of this study is to establish to what extent and in which ways PAUSE contributed to this control. Understanding the strategies that successfully worked to curb a daily rate grater than 11K in April to a daily rate of less than 1K in June are crucial in controlling potential resurgence of infection, and in guiding our efforts to prevent a new epidemic wave.

**Figure 6:**
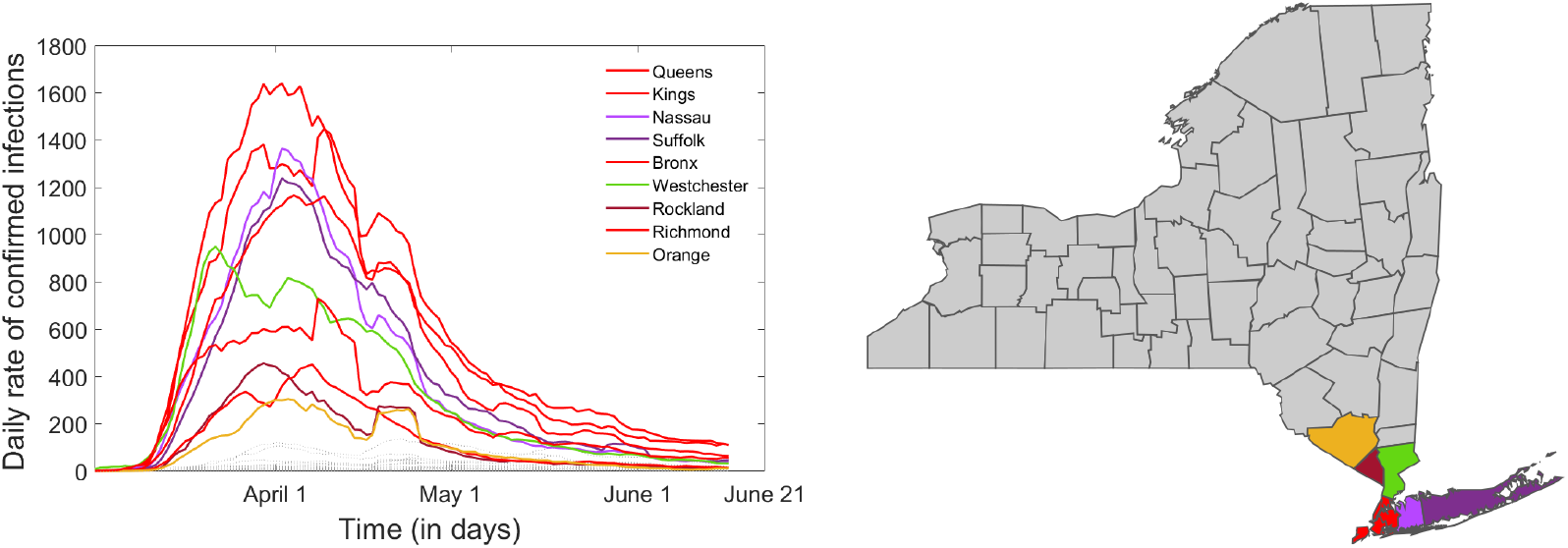
Daily infection rates in New York counties. form May 1st until June 20th. The counties with the highest daily rates of confirmed infections by June 20th are labeled and emphasized as solid curves. The rest of the counties, with lower infection counts, are shown as faded dotted lines, for comparison.

### 3.2 Traffic trends

The Apple driving time courses are highly correlated across New York counties (mean pairwise Searman coefficient 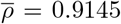 and mean significance *p* = 7.17 × 10^*−*7^). In broad strokes, they all show a sudden drop from baseline, initiated around March 4th (a few days after the first infections were confirmed in New York State), reaching an inflection point around March 12th (the day of the first government-mandated state-wide closures), dropping to a minimum value around March 22nd (the day PAUSE was instated), then slowly recovering towards the baseline (or even past the baseline, in some cases), at various upward rates (depending on the county). To illustrate these common trends, Figure 7 shows simultaneously the smoothed driving time courses for all 61 counties in New York State. Figure 8a shows the histogram of the deepest traffic drop from baseline. Figure 8b shows the histogram of the specific times when this minimum was achieved (all within a relatively narrow window around March 22nd). Figure 8c shows the histogram of the recovery end values on June 14th.

**Figure 7:**
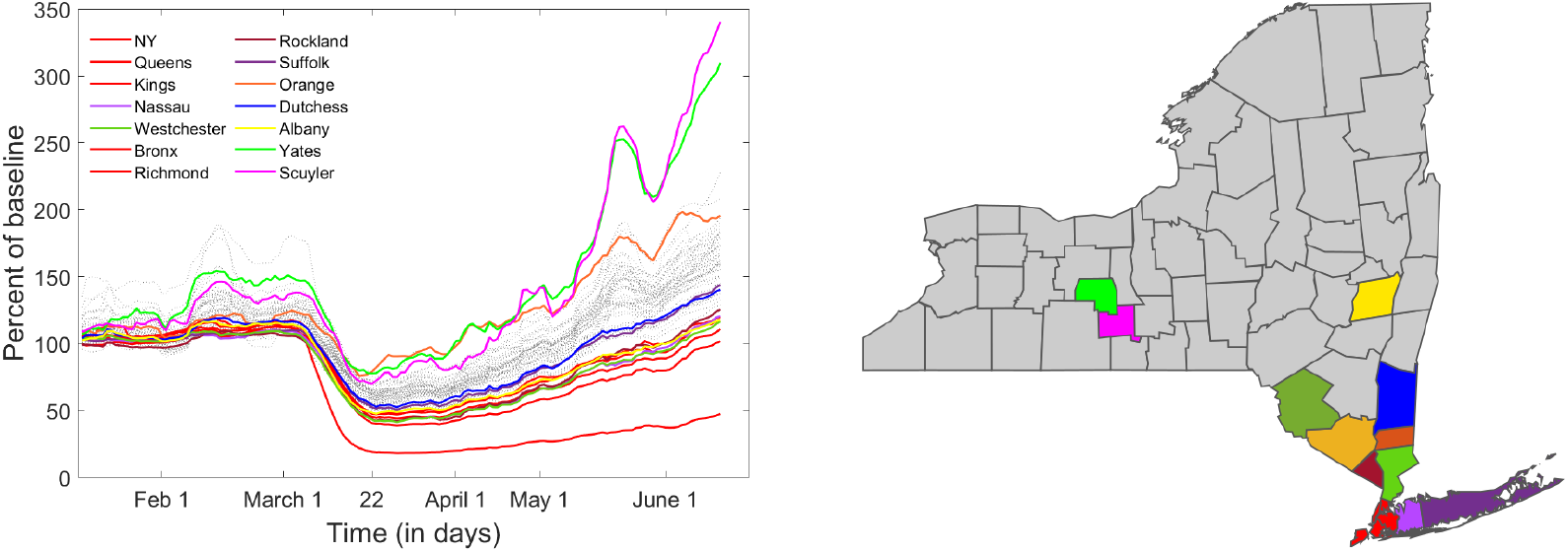
Description of driving trends in New York counties. **A**. Time course (of numbers of destinations per day with respect to baseline). Each curve shows the timeline for one county: the counties with high infection rates are color coded and shown in the legend; the others are shown as dotted grey lines. **B**. Each county’s location is shown in the corresponding color on the NYS map.

**Figure 8:**
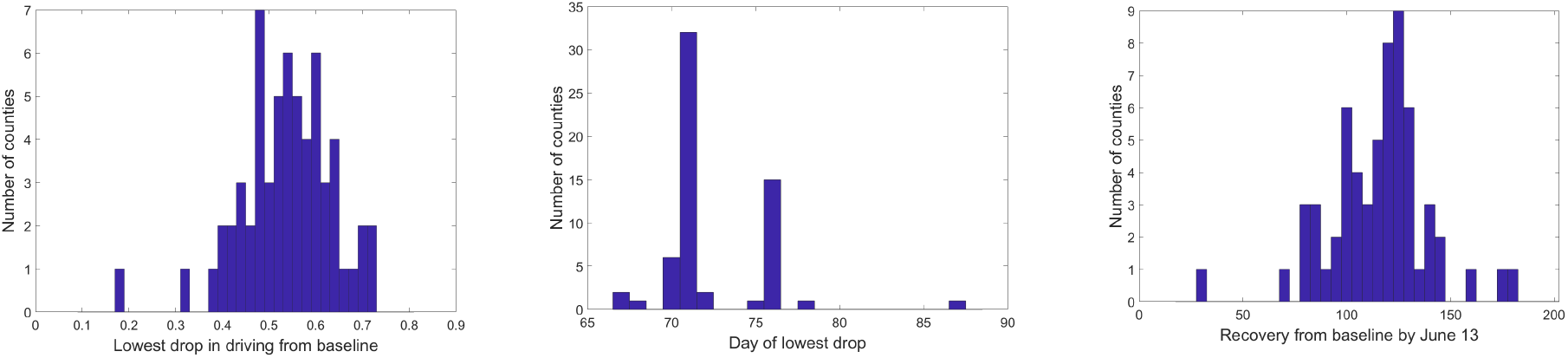
Description of driving trends in New York counties. **A**. Histogram of the values at the deepest drop in driving from baseline. **B**. Histogram of the day in the time course when the deepest drop occurred for each county. The few counties achieved their minimum after more than 75 days had prior close to minimum values. **C**. Histogram of the recovery driving level for each county on the last day computed by our smooth window averaging (June 12).

Broadly speaking, all NYS counties shows a highly coordinated drop in driving over an approximately two week long time window in March, due to the effect of PAUSE closing many places of business (and subsequently the traffic to and from them), modulated by the psychological and citizenship response to the increase in epidemic awareness and to the state directed measures.

However, there are also sizeable differences between the traffic timelines in each county, in particular in their path to recovery from this dramatic drop. While some counties (in particular those with highest COVID infection, among which the New York City counties) show a slower recovery rate, others have already recovered to the point of transcending the established baseline (which is not surprising, since the seasonal level of functional traffic for May/June is expected to be higher than the January baseline).

The differences in how social dynamics in New York regions responded to the epidemic is also reflected in different traffic patterns for the five metropolitan areas where information was provided on all three modalities (driving, walking and public transport). Figure 9 illustrates how different modalities recovered at different rates for the same city, and how these signatures differ between the five cities. Notice that the reduction and recovery in driving is relatively similar in all five cases (with the lowest point around the initiation of PAUSE and a recovery to 100% or slightly above baseline by June 19). The public transit suffered the most pronounced drop in all of the five cases, but the level of this drop and the recovery slope still differs between counties (with New York City showing, unsurprisingly, the most pronounced drop for a very high baseline of public transit, and also showing the slowest recovery (to less than 40% baseline by June 19). The most pronounced differences across these five areas are in walking patterns. Indeed, waking in the New York City shows a very slow recovery to baseline (slower than that of driving), as opposed to Rochester or Buffalo, where walking quickly recovered to values much higher than the baseline (which would be expected in absence of the epidemic, due to the seasonal increase with respect to the January-based baseline). This is not surprising, since the amount of walking is the most likely to be influenced under the current circumstances by factors like population density, geographic position (proximity to nature, and other large spaces which permit social distancing while walking). Unfortunately, a more in depth, county-wise analysis of walking patterns is not possible, since data is only provided for these five metropolitan areas.

**Figure 9:**
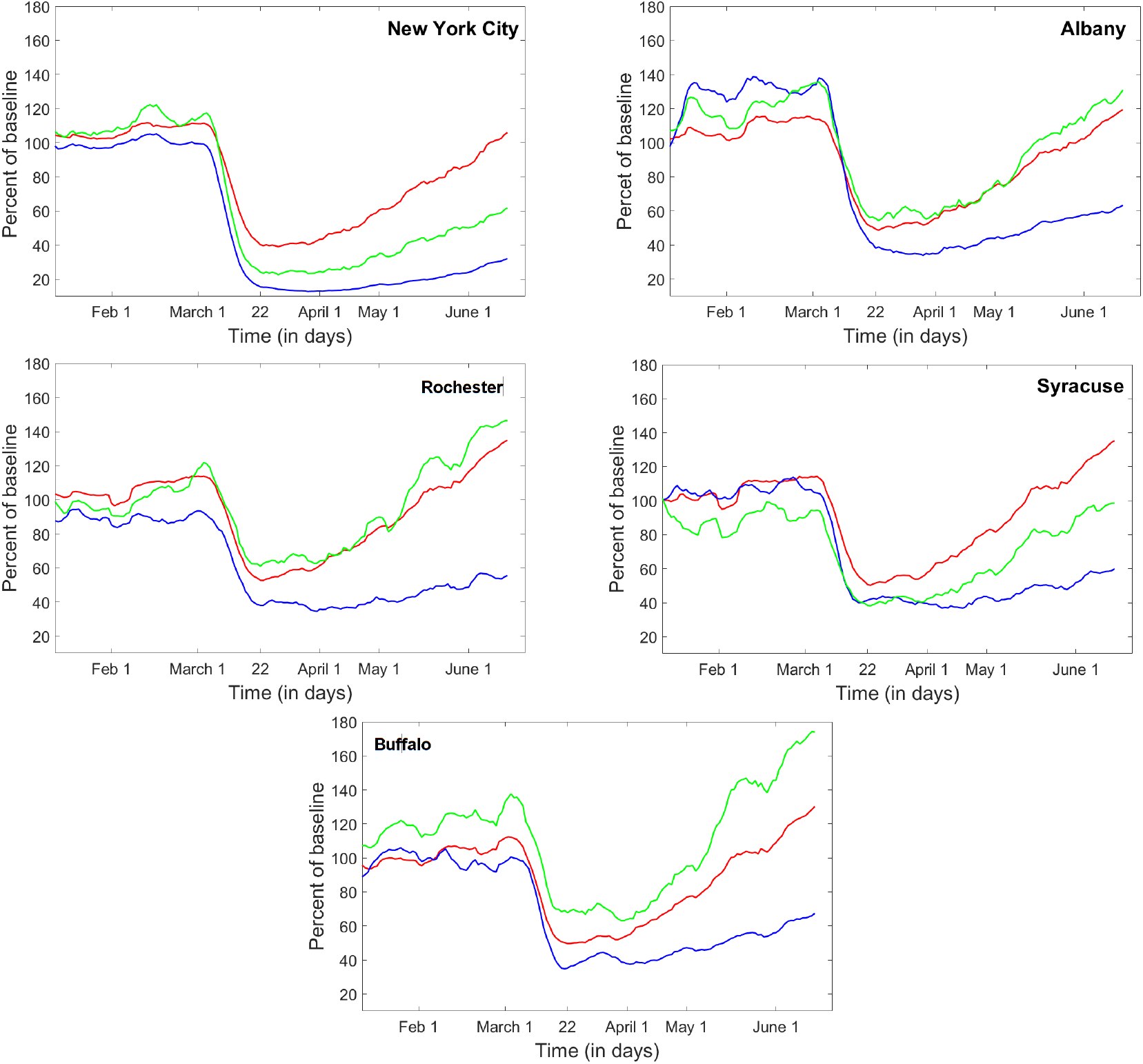
Trends in all traffic modalities (driving, walking and public transport) as provided for five metropolitan areas in New York State. **A**. New York City; **B**. Albany; **C**. Rochester; **D**. Syracuse; **E**. Buffalo. Driving patterns are shown as a red curve, walking in green and public transport in blue.

### 3.3 Location specific mobility trends

Similarly with the traffic timeline, mobility to the five location categories (Retail, Groceries, Parks, Transit and Workspace) was highly synchronized across New York counties, and so was the time duration spent at the residence.

Figure 10 illustrates simultaneously for all counties the mobility trends to all destinations, as well as the residential time. Each panel represents a destination, and each curve corresponds to the smoothed timeline in a different county. New York City counties (emphasized as solid lines in each panel) were in each case among the counties with the most pronounced effect: deep drop in mobility and high rise in residential time at PAUSE, followed by slow return towards the original baseline afterwards. Recall that New York City counties were also the ones to exhibit the most pronounced infection rates at PAUSE. The same trends were identified when using the weekly representation for mobility to destination and for residential times, as shown in Figure 11. In the next section, we will investigate whether this is a correlation between mobility and infection across all counties, and not just an isolated, or unique phenomenon specific to New York City. In order to avoid placing weight on the New York City counties (which are are the high end of the spectrum in both mobility and infection measures), we used a nonparametric, rank correlation.

**Figure 10:**
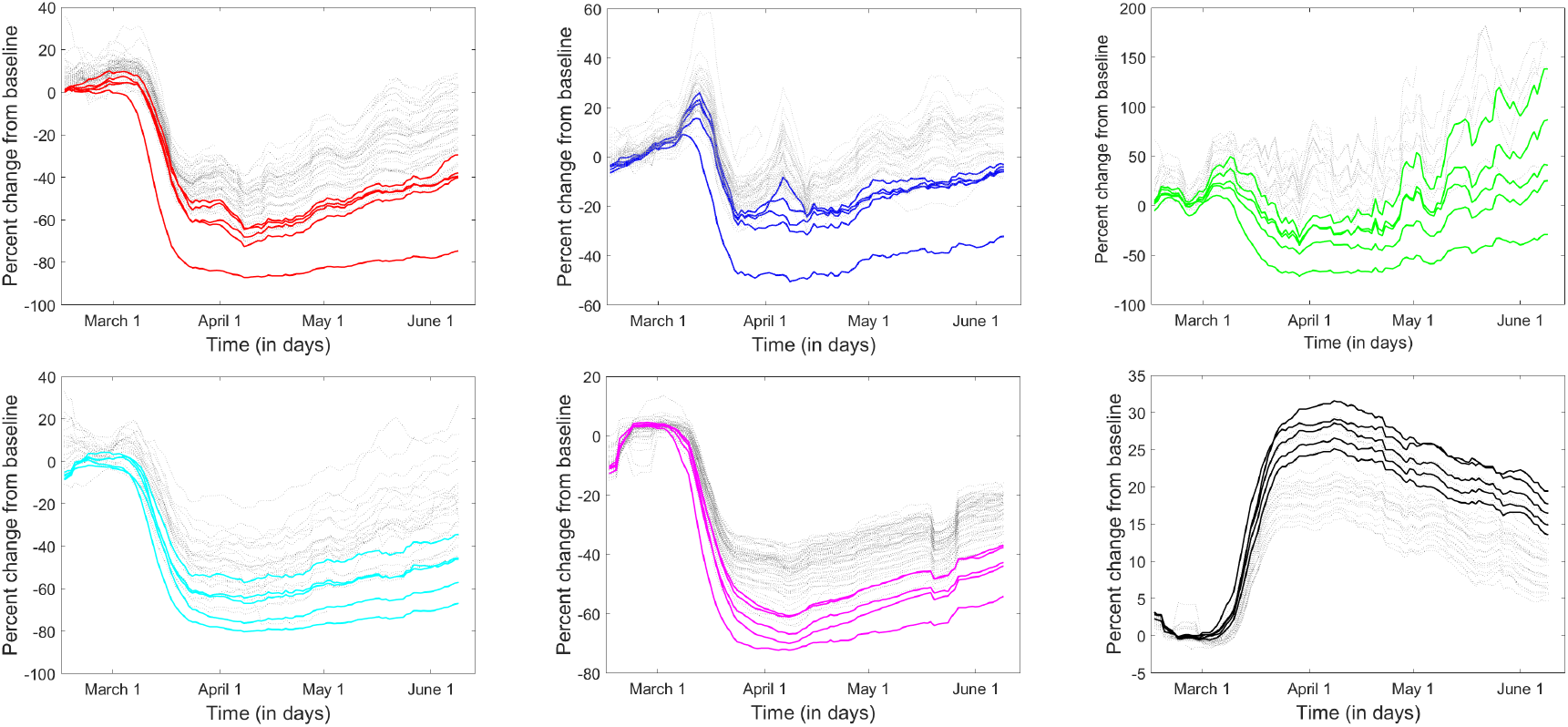
Seven day moving window average time series describing mobility to **A**. Retail and Recreation; **B**. Grocery and Pharmacy; **C**. Parks; **D**. Public Transit; **E**. Workspace; **F**. Residential time. New York City counties are shown in thick solid lines; all other counties are shown in grey dotted lines.

**Figure 11:**
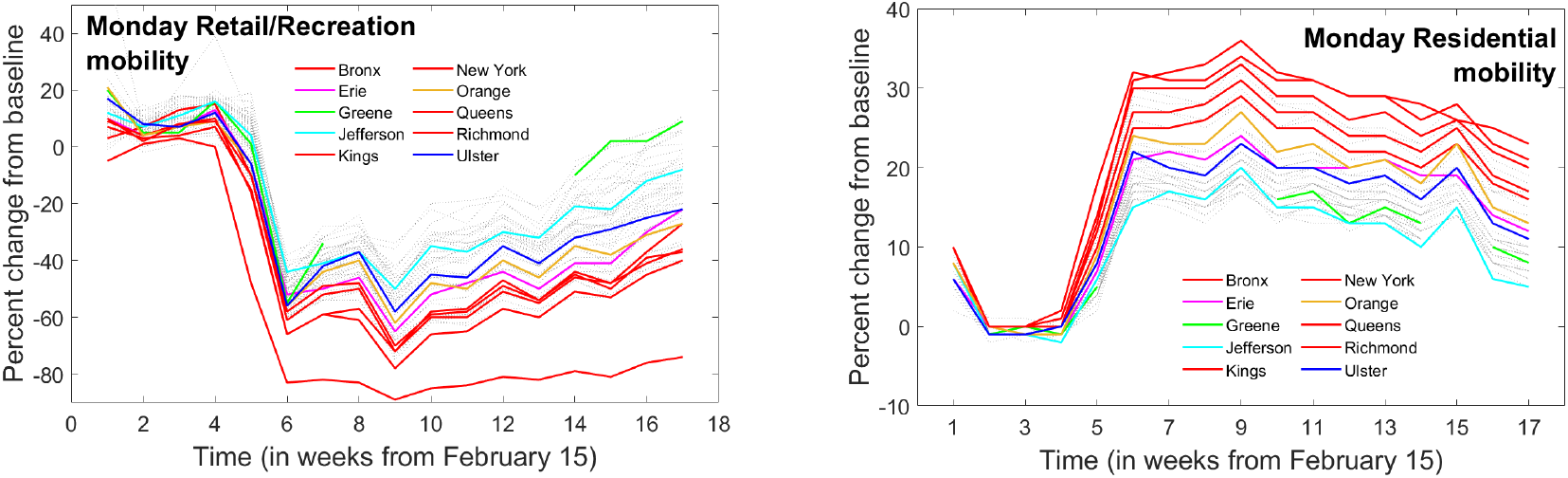
Weekly Monday mobility trends for all counties, for **A**. Retail and Recreation and **B**. Residential time.

Another broad and hardly surprising observation is that the trips to retail, work and transit dropped significantly lower than those to groceries and parks. Park and the outdoors varied widely between counties; as we mentioned before, this is likely based on the geographic position and access to nature (large outdoor spaces where social distance can be observed). These observations are supported by Figure 12, which compares mobility patterns for all six compartments between three example counties, one in each infection cluster from Figure 5: New York County, Suffolk County and Ulster County. In New York County, for example, mobility to Parks decreased to a comparable extent to mobility to Retail, Transit and Workspace destinations, after which it started a very slow recovery, and remained below the January baseline, even by June 19. In contrast, mobility to Parks in Suffolk and Ulster Counties shows an increasingly rising trend from baseline, seemingly uncorrelated to the PAUSE date.

**Figure 12:**
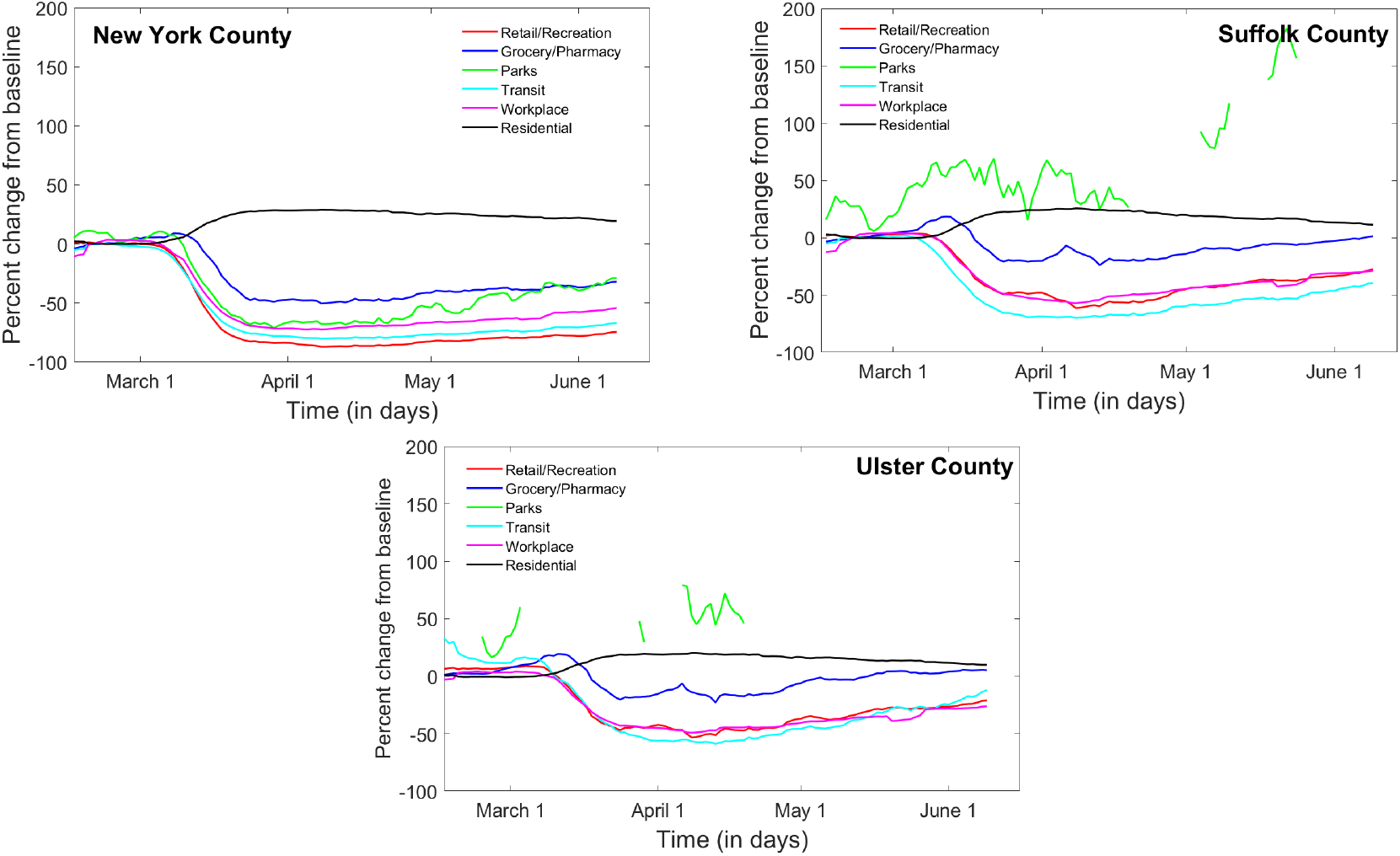
Seven day moving window average time series for mobility to all destinations for **A**. New York County; **B**. Suffolk County; **C**. Ulster County.

Information on mobility to Parks would be of particular interest, in the context of using the data to assess infection risk between outdoor recreational and other activities. Unfortunately, the Parks time series have many gaps in reporting, which are augmented by the windowed smoothing. Further processing the data to account for these gaps would render any assessment of intrinsic trends unreliable.

### 3.4 Effects of infection on traffic and mobility

One direction of inquiry of our study is to establish whether the size of the epidemic had any effect on the efficiency of the lock down measures. To explore this, we considered a few potential measures of the outbreak size: (1) number of confirmed infections, (2) incidence of confirm infections versus total tests, and (3) rate of confirmed infections on the day PAUSE was initiated (May 22). As measures of the PAUSE efficiency, we considered traffic and mobility activity at their lowest level (which we showed to be highly synchronized in time with PAUSE, across counties, modalities and venues). Because of the missing data in some of the mobility timelines (primarily due to Google privacy thresholds), we could only sustainably calculate the lowest point for the Retail, Groceries and Workspace time series. While the most variability between counties appears to occur in travel to outdoor Parks, that is also the data which had the largest gaps, making a consistent analysis impossible.

All measures of epidemic size correlated negatively with the mobility level; in other words, the higher the infection (be it measured in sheer numbers, incidence rate per tests, or daily infection rates) the more severe was the drop in activity. This proved to be true for both driving trends (shown in the first row of Table 1), and for mobility to Retail, Groceries and Work (shown in rows 2-4 of Table 1).

**Table 1:**
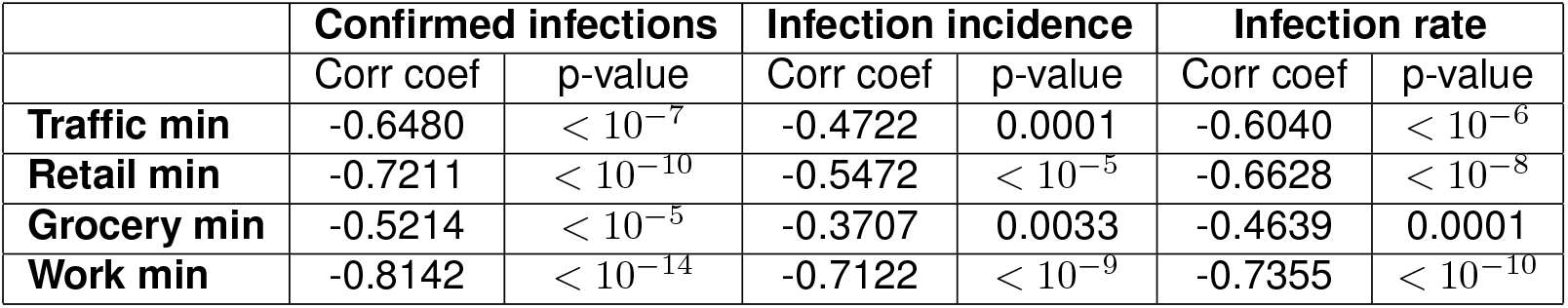
Spearman correlations between measures of epidemic size and measures of social mobility in New York counties at the time of PAUSE. The epidemic measures used are: number of confirmed infections (two left columns); number of confirmed infections per number of tests in each county (two middle columns); daily infection rate for each county (two right columns). These were correlated with the lowest traffic level, as fraction of traffic baseline (top row), and with the lowest mobility levels to Retail, Grocery and Workspace (remaining three rows). The corresponding significance values are shown as separate columns.

As we had mentioned in the Methods section, the Google mobility data is best used and interpreted separately for each week day (since different week days have different baseline values, hence it is problematic to mix, average or compare the baseline percent changes with each other). Therefore, in order to increase the confidence of our correlations, we also did the statistical analysis using week day specific time series.

Table 2 illustrates the correlation values for the Tuesday and Sunday mobility data. The other week days were similar, and are not shown. Notice that Retail and Work correlated most strongly with the epidemic measures. Traffic (a less reliable measure in terms of destination and exposure) showed weaker correlations, and travel to Groceries (more of a daily necessity) showed the weakest correlations. Also, the correlations were stronger for the “working” week days than for the weekend days.

**Table 2:** Spearman correlations between measures of epidemic size and measures of social mobility in New York counties at the time of PAUSE. The epidemic measures are: number of confirmed infections (two left columns); number of confirmed infections per number of tests in each county (two middle columns); daily infection rate for each county (two right columns). The lowest mobility levels to Retail, Grocery and Workspace were computed separately for each day of the week; the table shows the results for correlations with Tuesday and Sunday time series.

|  | Tuesday |  |  |  |  |  |
| --- | --- | --- | --- | --- | --- | --- |
|  | Confirmed infections |  | Infection incidence |  | Infection rate |  |
|  | Corr coef | p-value | Corr coef | p-value | Corr coef | p-value |
| **Retail min** | -0.7102 | $< 10^{-9}$ | -0.5913 | $< 10^{-6}$ | -0.6350 | $< 10^{-7}$ |
| **Grocery min** | -0.4693 | 0.0001 | -0.4591 | 0.0001 | -0.4920 | $< 10^{-4}$ |
| **Work min** | -0.8274 | $< 10^{-15}$ | -0.7047 | $< 10^{-9}$ | -0.7355 | $< 10^{-10}$ |
|  | Sunday |  |  |  |  |  |
|  | Confirmed infections |  | Infection incidence |  | Infection rate |  |
|  | Corr coef | p-value | Corr coef | p-value | Corr coef | p-value |
| **Retail min** | -0.5323 | $< 10^{-4}$ | -0.3415 | 0.0071 | -0.4543 | 0.0002 |
| **Grocery min** | -0.2545 | 0.0478 | — | — | — | — |
| **Work min** | -0.7166 | $< 10^{-10}$ | -0.5776 | $< 10^{-5}$ | -0.6081 | $< 10^{-6}$ |

Altogether, this first correlation test shows that the degree to which social mobility was restricted was more pronounced in counties with higher infection. The correlation was stronger for destinations which did not represent daily necessities, and less pronounced for destination which were either essential (food, medicine) or safer (outdoors). These ideas will be revisited as a discussion point in Section 4.

### 3.5 Effects of mobility restrictions on controlling the epidemic

Once we established a correlation between the infection count and the size of the traffic / mobility drop, the next important question is to investigate whether the mobility drop efficiently acted to slow down transmission. We did so by computing correlations between the traffic / mobility drop at PAUSE, and two measures of epidemic control: increase in confirmed cases, and change in daily rate (from May 22 until June 21).

Table 3 shows the results of this correlation analysis for driving trends, as well as for smoothed time series corresponding to mobility to Retail, Groceries and Workspace. Table4 shows the correlation values when using week day based mobility time series to Retail, Groceries and Workspace.

**Table 3:** Spearman correlations between social mobility restriction in New York counties at the time of PAUSE, and epidemic control by June 20th. As measures of epidemic control, we used the rise in confirmed infections, and the drop in infection daily rate (between March 22 and the last value in our smooth time series). These were correlated with the lowest traffic level, as fraction of traffic baseline (top row), and with the lowest mobility levels to Retail, Grocery and Workspace (remaining three rows). The corresponding significance values are shown as separate columns.

|  | Rise in confirmed infections |  | Drop in infection rate |  |
| --- | --- | --- | --- | --- |
|  | Corr coef | p-value | Corr coef | p-value |
| **Traffic min** | -0.4454 | $< 10^{-6}$ | -0.0002 | $< 10^{-4}$ |
| **Retail min** | -0.6329 | $< 10^{-7}$ | -0.4563 | 0.0002 |
| **Grocery min** | -0.4064 | 0.0012 | -0.2994 | 0.0191 |
| **Work min** | -0.7885 | $< 10^{-13}$ | -0.5083 | $< 10^{-4}$ |

**Table 4:** Spearman correlations between social mobility restriction in New York counties at the time of PAUSE, and epidemic control by June 20th. As measures of epidemic control, we used the rise in confirmed infections, and the drop in infection daily rate (between March 22 and the last value in our smoothed time series). The lowest mobility levels to Retail, Grocery and Workspace were computed separately for each day of the week; the table shows the results for correlations with Tuesday and Sunday time series.

|  | Tuesday |  |  |  |
| --- | --- | --- | --- | --- |
|  | Rise in confirmed infections |  | Drop in infection rate |  |
|  | Corr coef | p-value | Corr coef | p-value |
| **Retail min** | -0.5892 | $< 10^{-6}$ | -0.4801 | $< 10^{-4}$ |
| **Grocery min** | -0.3286 | 0.0097 | -0.4032 | 0.0013 |
| **Work min** | -0.7780 | $< 10^{-12}$ | -0.4853 | $< 10^{-4}$ |
|  | Sunday |  |  |  |
|  | Rise in confirmed infections |  | Drop in infection rate |  |
|  | Corr coef | p-value | Corr coef | p-value |
| **Retail min** | -0.4913 | $< 10^{-4}$ | -0.2184 | 0.0909 |
| **Grocery min** | -0.2130 | 0.0993 | — | — |
| **Work min** | -0.6818 | $< 10^{-8}$ | -0.4869 | $< 10^{-4}$ |

Our analysis shows that deeper traffic and mobility drop correlated with better epidemic control, as measured by the progress in epidemic recovery (“flattening the curve”) from PAUSE (March 22) until the date of our study (June 20). When PAUSE was initiated, it was first suggested that it its first effects on flattening the infection curve would be visible within 14 days (which is the approximate incubation period of the COVID virus, and the mean time window from exposure to symptom development and likely testing and diagnosis). We would like to investigate whether the data suggests a better estimate of how far after initiating PAUSE the curve flattening started to take effect.

To accomplish this, we calculated the progression of the correlation between the mobility drop and the epidemic recovery (estimated using the same measures as before), but with a rolling end date from March 23 (just after initiation of PAUSE) until June 20 (the date of our study). Figure 13 shows how the correlation coefficients (and significance value) change with respect to the rolling end date.

**Figure 13:**
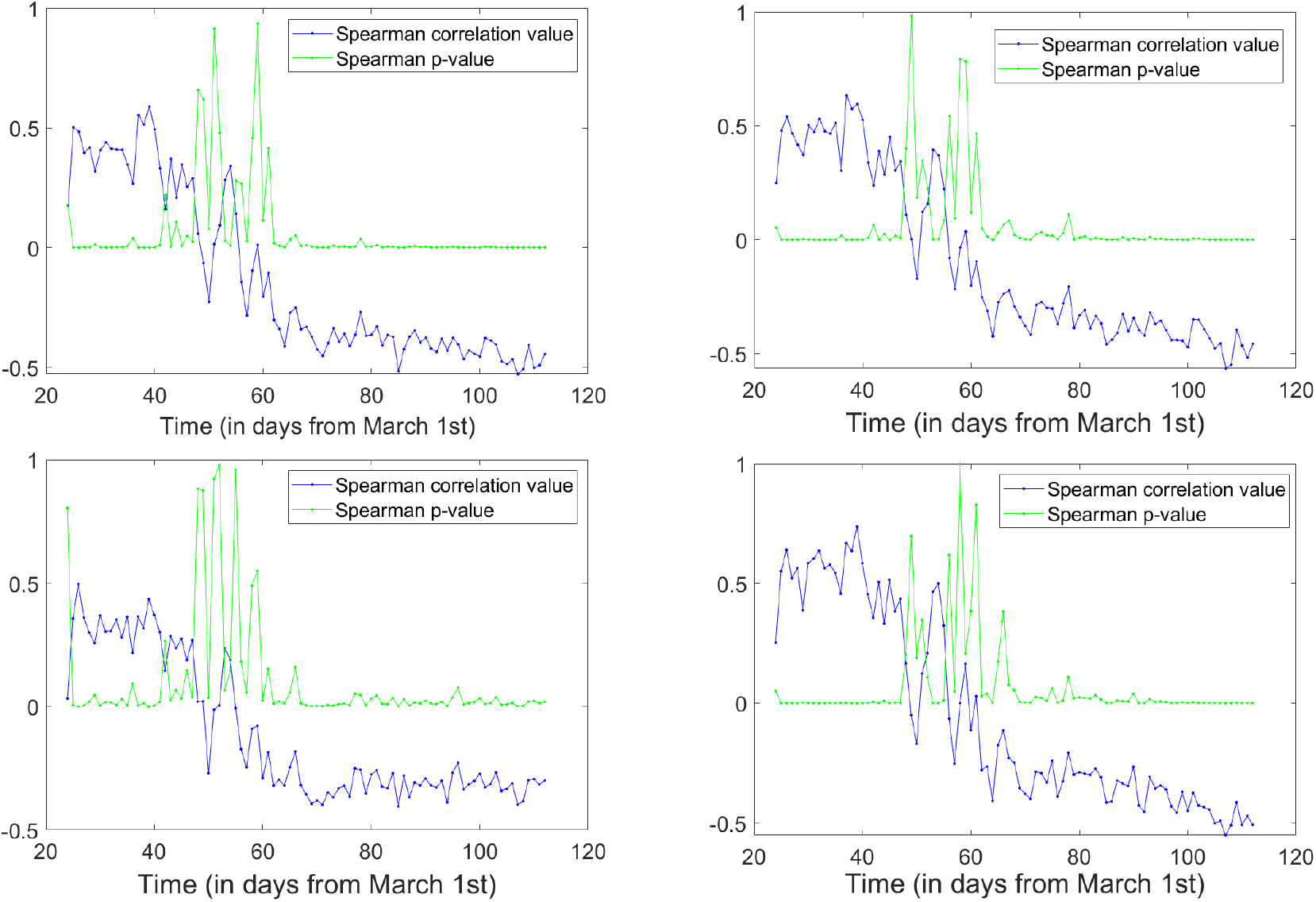
Correlations between traffic drop at PAUSE and efficiency of epidemic control. For each day after PAUSE (represented along the x-axis) we correlate the drop in daily infections slope compared to that at PAUSE, with the lowest drop in **A**. driving, as percent of baseline; **B**. mobility to Retail, **C**. mobility to Grocery, **D**. mobility to Workspace, as percents of the reference day. The Spearman correlation values are shown in blue for each day, and the significance p-values are shown in green.

All representations in Figure 13 are phenomenologically similar, in the sense that the lowest drop level in both traffic and mobility shows a significant *positive* correlation with the increase in epidemic rate for the first window of about 20 days. This is followed by a transition window in which there is no consistent significant correlation. Finally, after about 40 days from PAUSE, the variables start to show a significant negative correlation, consistently to the end of the time interval (the final values are the ones shown in Table 3). The graphs suggest that, for a while, the epidemic rate kept increasing faster for deeper drops in traffic. This is not a surprise, since we showed that deeper traffic drops were symptomatic of high infection rates at PAUSE, which are expected to take a little while to be curbed. Indeed, this appears to be the case: after 5-6 weeks from the initiation of the shutdown, deeper PAUSE drops in traffic are correlated consistently larger reductions in daily rates (at this point most counties have passed their peak rate). This is important, since it may be indicative of the fact that PAUSE took about 5-6 weeks to show consistent and measurable results, and will be further discussed in our last section.

### 3.6 Effects of mobility recovery on controlling the epidemic

As Figures 7 and 10 suggest, both traffic and mobility recovered from PAUSE at different rates and to different extents across New York counties. As mentioned before, it is likely that the increase in mobility has much to do with the density, location, and current epidemic reports in each county. Our last question addresses how mobility recovery measures are coupled with epidemic recovery measures. A strong tool in establishing this relationship is calculating cross-correlations between the two types of time series, and infer which one is likely to drive the other. Unfortunately, the frequent gaps in our mobility time series did not permit this longitudinal analysis, which we were only able to perform for the driving data).

Separately for each county, and for each day after PAUSE, we calculated the traffic recovery (as percent change from baseline) from its lowest drop (at PAUSE). We then correlated it with the epidemic recovery (measured as the drop in daily infection rates between the value at PAUSE and the value on the day in question). Figure 14 shows the correlation value and the significance value for each day. We observe the same pattern, consisting of a window of about 20 days after PAUSE where the two measures are positively correlated, followed by a transit in which there is no significant correlation; starting 5-6 weeks after PAUSE, the two measures show a consistent and significant negative correlation: faster return to baseline driving based activities correlates inversely with the efficiency in curbing the epidemic rate.

**Figure 14:**
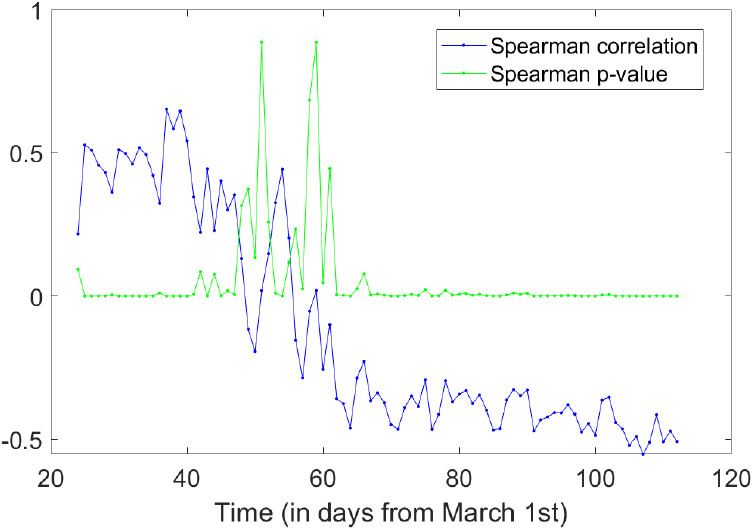
Correlations between driving recovery from PAUSE and efficiency of epidemic control. For each day after PAUSE (represented along the x-axis) we correlate the difference in driving levels between the current day and PAUSE (as percent of baseline) with the difference in daily infections slopes between PAUSE and the current day. The Spearman correlation values are shown in blue for each day, and the significance p-values are shown in green.

## 4 Discussion

Our study investigated epidemic and social traffic patterns in New York State counties since the advent of COVID 19 in March 2020, until June 21, 2020. We first focused on identifying the signature of the time series in each data set separately, and on drawing qualitative and quantitative comparisons between the situation in each county. We then aimed to understand the dynamics interplay between the two, by testing for correlations between measures describing (1) size, timeline and control of the epidemic, and (2) the reduction in social traffic due to PAUSE directives, and the subsequent relaxation process.

Our first analysis found the number of infections in each county to be significantly negative correlated with the level to which the social traffic dropped in each county at the start of PAUSE. This correlation persisted when we considered daily infection rates, or incidence rates as an alternative measure of the epidemic. This supports the idea that psychology played a crucial role in the shutdown process, and that counties with reportedly higher risk were able to enforce a more efficient social distancing protocol.

Conversely, our second analysis found significantly negative correlations between the level to which traffic dropped in each county, and the reduction in epidemic rate post PAUSE. This supports the idea that PAUSE had a large part to play in the process of curbing the infection rate, but also suggests a secondary mechanism: counties with high infection rates were more motivated to act more drastically and faithfully along the PAUSE directives, which lead to deeper cuts in infection. We noticed that the curve flattening became significant to our correlation statistics around 40 days after the start of PAUSE, which is longer than the 14 day period that was a popular expectation with the general public, based on the incubation period of the virus. Our study suggests that a longer period is needed in order to allow sizeable effects to emerge in the population.

Finally, our last analysis found significant negative correlations between the level to which driving patterns recovered in each county since the start of PAUSE, and the reduction in epidemic rate. This suggests that a slower return to normal activities may have facilitated the slowing down of infection.

Altogether, these facts support not only the importance of the PAUSE directive, but also of the degree and length to which it was observed. Our results, in combination, emphasize the efficiency of social distancing measures, and the importance of understanding their necessity.

## Data Availability

All data in the manuscript was taken from the public domain. The sources are references in the bibliography.

## Acknowledgements

This work was supported by the SUNY New Palyz AC^2^ Summer Research Program. We would like to thank Dr. Nancy Campos, for her constant and invaluable input on our work.

